# The Effect of Residents’ Working-Hour Restrictions on Patient Safety, Resident Well-Being, and Resident Education: Protocol for a Systematic Review and Meta-Analysis

**DOI:** 10.1101/2021.08.26.21262689

**Authors:** Gabriela Zavala Wong, Maitza Vidal Meza, Maria Lazo-Porras

## Abstract

**Introduction:** Residents’ duty hour restrictions have been a source of debate throughout the years, given that extended shifts have historically been associated with a negative effect on patient safety. Implementing restricted duty hours may help reduce sleep deprivation and workload, consequently improving residents’ sense of well-being. On the other hand, these reforms implicate a greater number of handoffs where communication errors may arise, and continuity of care being lost as a result. In a similar way, shorter shifts may implicate less time of direct patient contact and, consequently, decreased educational opportunities for residents. Various studies have attempted to explore the effect of resident work hour reforms on patient safety outcomes. However, these have been mainly based solely on observational studies that have not been subjected to the same rigor as experimental ones, primarily because no randomized controlled trials (RCT) were available in this matter. Nonetheless, more substantial evidence has become available in these last few years as three RCTs have been published exploring the impact of resident duty hour restrictions on patient safety as well as on residents’ wellbeing and education. An updated systematic review and meta-analysis are crucial to interpret this data that has now become available.

**Objectives:** To evaluate the effect of resident physicians’ working-hour restrictions on patient safety parameters, residents’ perceived well-being and resident education.

**Methods and analysis:** This research protocol was developed according to PRISMA-P and the Cochrane guidelines for systematic reviews and metanalysis. Electronic literature search strategies were developed using MeSH and free terms to be carried out in PubMed, MEDLINE, EMBASE, Cochrane Library, Clinicaltrials.gov and Global Index Medicus with no restriction in language. Primary outcome measures include several patient safety parameters, whereas secondary outcome measures involve resident well-being and education. Two research team members will screen identified titles, abstract and full text, evaluate risk of bias and extract data in an independent manner. A qualitative narrative synthesis will be employed to summarize the key findings, population, and methodology of studies using text and tables for both primary and secondary outcomes. We will test for heterogeneity of the included studies by employing the I^2^ statistical test; if significant (I^2^ ≥ 75%), only qualitative synthesis will be presented. On the contrary, if studies are homogeneous, a meta-analysis will be considered using Review Manager 5.1 software. For continuous data, we will calculate the mean difference or standardized mean difference. For dichotomous data, the risk ratio (RR) will be calculated. Results will be displayed on a Forest Plot. To assess bias, a Funnel plot and Egger test will be employed.

**Conclusions:** This systematic review will provide evidence regarding the effect of resident physicians’ working-hour restrictions on patient safety parameters, residents’ perceived well-being and resident education. All of these are variables that must be considered when determining policies regarding the medical training environment. It is essential to review the existing high-grade evidence regarding the impact of residents’ extended working hours so that authorities can optimize future graduate medical education regulations.

## INTRODUCTION

Prolonged hospital shifts have historically been a distinctive characteristic of medical residency programs. However, regulations on these working conditions have been a subject of constant debate over the years. In 2003, the US Accreditation Council for Graduate Medical Education (ACGME) announced reforms in duty hours. Under these new regulations, residents were to work no more than 80 hours per week, have shifts no longer than 24 hours, and have at least 10 hours of rest between shifts (1)]. These reforms were supported by studies that demonstrated that sleep deprivation harms residents’ well-being and a fatigued staff is more likely to commit medical errors(1,2). Under this same argument, later in 2011, the ACGME further reduced interns’ shifts to 16 consecutive hours and 28 hours for senior residents(2). Nevertheless, new studies allege that hour restrictions do not affect patient safety and therefore, ACGME, in 2017, updated regulations to allow for longer shifts for interns (3), but to this date there is no international consensus on the maximum limit of workload for medical residents.

There is controversy regarding the length of hospital shifts and adequate rest for safe medical practice. On the one hand, extended shifts might have a detrimental effect on patient safety by interfering with physicians’ mental and physical health (4). Previous studies have shown that, indeed, alertness and performance vary according to a person’s circadian rhythm. Sleep deprivation, along with a high workload, could impair a person’s ability for diagnostic problem solving and complex treatment decision-making (5,6). Under this pretense, a well-rested resident physician would provide safer care to patients and consolidate knowledge better and acquire greater clinical skills for their future independent practice. On the other hand, however, shorter shifts implicate a more significant amount of patient handoffs, which may pose an even greater threat to patient safety given that handover-related communication errors may arise, and continuity of care may be lost as a result (7). In a similar way, shorter shifts may implicate less time of direct patient contact and, consequently, decreased educational opportunities.

Various studies have attempted to explore the effect of resident work hour reforms on patient safety outcomes. The most recent systematic reviews have concluded that these restrictions had no impact on patient care indicators (8,9). However, these have been mainly based solely on observational studies that have not been subjected to the same rigor as experimental ones, primarily because no randomized controlled trials (RCT) were available in this matter. Nonetheless, more substantial evidence has become available in these last few years as three RCTs have been published exploring the impact of resident duty hour restrictions on patient safety. The Flexibility in Duty Hour Requirements for Surgical Trainees (FIRST) trial, which ran from July 2014 to June 2015, randomly assigned 119 surgical residency programs in the United States to adhere to either standard ACGME duty-hour rules or flexible schedules that waived shift length limits and resting time between shifts (10). Likewise, the Individualized Comparative Effectiveness of Models Optimizing Patient Safety and Resident Education (iCOMPARE) trial, carried out from July 2015 to June 2016, included 63 internal medical residency programs and explored similar outcomes (11). The most recent Randomized Order Safety Trial Evaluating Resident-Physician Schedules (ROSTERS), which ran from July 2013 to March 2017, investigated the effect of eliminating extended shifts on patient safety in six pediatric intensive units across the US (12). An updated systematic review and meta-analysis are crucial to interpret this data that has now become available. Additionally, the studies mentioned above have also explored multiple other outcomes simultaneously, including the impact of resident duty hour restrictions on residents themselves, particularly their well-being and education. All of these are variables that must be considered when determining policies regarding the medical training environment.

## JUSTIFICATION

Residents are an essential component of the medical workforce, particularly in low- and middle-income countries (LMIC), where healthcare providers have a significant shortfall in the public sector. In this context, resident doctors are mainly responsible for the effective implementation of quality health care services that would not otherwise meet the needs of the population. However, while trying to bridge this labor shortage, authorities often overlook that residency is an educational system whose primary purpose is to optimally prepare the physician for their future independent practice as a medical specialist. To achieve this, they must provide adequate training conditions. In Peru, despite the heterogeneity of regulations across hospital teaching centers, previous studies have shown that residents identify several issues related to working conditions and infrastructure (13–15). Among the most frequent complaints are excessive workload and extended shifts that develop into anxiety, frustration, and ultimately resident burnout; the risk being that any shortcoming in a resident’s performance may affect a patient (13). Similarly, residents perceive that learning objectives are not always fulfilled under these training conditions (14). As such, it is essential to review the existing high-grade evidence regarding how extended working hours influence patient safety outcomes, residents’ well-being, and education so that authorities can optimize future graduate medical education regulations. Previous systematic reviews have shown that most experimental studies evaluating residency working hour regulations have been carried out in high-income countries (HIC). However, this particular review will consist of a global literature search to also identify studies carried out in LMICs in order to develop recommendations for residency program authorities that can be applied worldwide.

## OBJECTIVES/SPECIFIC AIMS

### Primary objective

- To assess the effect of resident physicians’ working-hour restrictions on patient safety parameters.

### Secondary objective

- To evaluate the impact of resident physician’s’ working-hour restrictions on residents’ perceived well-being.
- To evaluate the impact of resident physician’s’ working-hour restrictions on residents’ objective education outcomes and self-reported satisfaction with quality of education received.

## METHODS AND ANALYSIS

### Study design

This protocol was designed according to Preferred Reporting Items for Systematic Reviews and Meta-Analysis Protocols (PRISMA-P) (16). Published results will follow the PRISMA 2020 statement (17).

### Eligibility criteria

Studies will be selected according to the criteria outlined below.

#### Types of studies

For our primary objective, we will include all randomized controlled trials (RCTs) in resident physicians that compare the effectiveness of working-hours restrictions vs. traditional extended-duration work schedules on patient safety, where at least one of the following is reported: mortality within 30 days, readmission, adverse events, medical errors, serious medical errors, in-hospital complications, 30-day readmission rate, prolonged hospital length of stay and/or patient safety indicators. For our primary objective, we will exclude quasi-experimental studies (pre and post-intervention designs).

For secondary outcomes, we will include RCTs and quasi-experimental studies (pre and post-intervention designs). We will exclude case-control studies, case series, and case reports.

We will include all relevant studies regardless of the language in which they were published. For studies published in a language other than English or Spanish we will use an online translator.

In the case of studies that evaluate working-hour restrictions in healthcare professionals other than medical residents (e.g: attending physicians, physician assistants, nurses, dental professionals, technicians, therapists, pharmacists), they will be included only if the results are divided and distinguished by occupation. Therefore, only information for medical residents will be selected, excluding other healthcare professionals.

#### Participants

We will include studies examining resident physicians enrolled in residency programs with no restrictions on their year of training or medical specialty.

We will exclude studies carried out in other healthcare professionals including attending physicians, physician assistants, nurses, dental professionals, technicians, therapists, pharmacists.

#### Intervention

Of interest are interventions addressing working-hour restrictions. The definition of working-hour restrictions was considered as reducing shift length (the number of consecutive hours that a medical resident works without protected sleep) and/or implementing a night float system (where an assigned resident works exclusively during the evening/night) (2)We will not consider a minimum or maximum duration of the intervention. We will exclude studies that involve implementation of protected sleep periods, time-management tactics or wellness workshops as the intervention.

#### Comparison

A restricted schedule will be compared with traditional extended shifts that do not have a restriction on consecutive hour shift limits and/or do not implement a night float system.

#### Outcomes

The primary outcome will be based on patient safety parameters including:

● 30-day mortality
● 7-day readmission
● 30-day readmission rate
● Adverse events (18): undesirable injury caused by medical care. If possible these will be further classified into non-preventable (without apparent error) and preventable (non-intercepted error).
● Medical errors (19): Any error in the delivery of medical care. If possible, it will be further classified into:
  ○ Serious medical error (19): medical error that harms the patient or has substantial potential to cause injury.
  ○ Near miss (19): error in medical care that has substantial potential to harm the patient but does not.
  ○ Error with no potential for harm (19): error that is unlikely to harm a patient
● In-hospital complications
● Length of stay
● Patient safety indicators (20): based on provider-level indicators of the Agency for Healthcare Research and Quality (AHRQ).

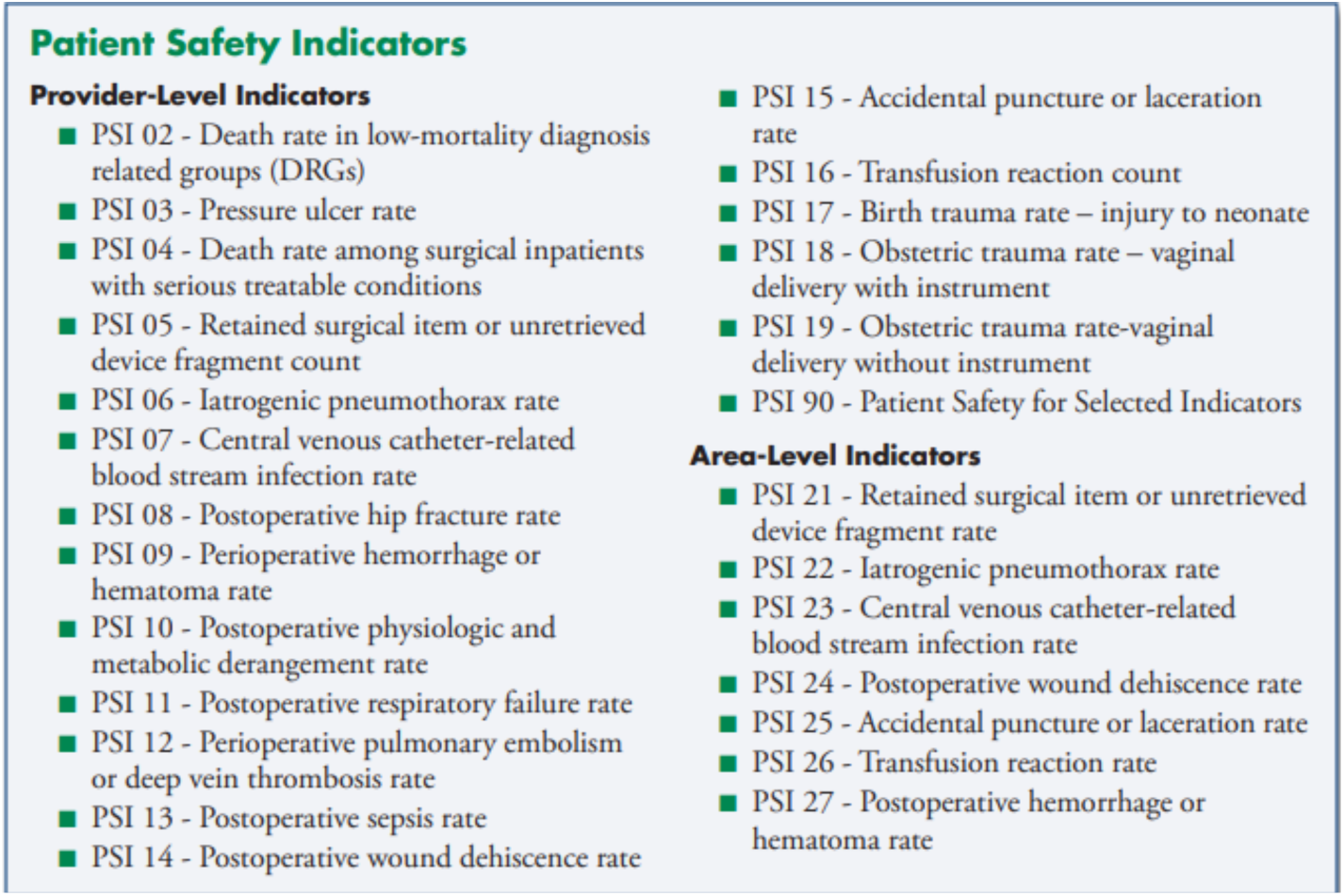

(Image from AHRQ Quality Indicators™ Patient Safety Indicators [Internet]. 2015 [cited 2021 Jun 14]. Available from: www.qualityindicators.ahrq.gov)

Secondary outcome measures will include:

#### Resident Education

● Resident’s examinations scores:
  ○ Performance on oral and written American Boards examinations
  ○ Performance on in-training examinations
● Resident’s self-reported satisfaction with the educational experience:
  ○ Rounds/teaching conference attendance
  ○ Independent reading/ self-directed learning
  ○ Miss OR/leave during operation
  ○ Resident autonomy in medical decision making
  ○ Continuity of care
  ○ Bedside teaching
  ○ Teaching satisfaction

#### Resident Well-Being

● Resident’s self-reported levels of rest, professional satisfaction, overall well-being, personal safety, fatigue and work/life balance, which include:
  ○ Professional satisfaction
    ■ Job
    ■ Working hours/schedule
    ■ Hands-off
  ○ Personal safety
    ■ Percutaneous/Attentional injuries
    ■ sleepiness
    ■ burnout
    ■ number of vehicle crashes
    ■ number of near vehicle crashes
  ○ Work/life balance:
    ■ Time for family/friends
    ■ Time for hobbies
    ■ Vacation
    ■ General Health
    ■ Bodily pain
    ■ Mental Health

#### Timing

No restriction will be applied for a year of publication.

#### Setting

There will be no restrictions by type of setting.

#### Language

No language restriction will be applied for any of the studies included for primary and secondary outcomes.

## INFORMATION SOURCES

Electronic literature search strategies will be developed using medical subject headings (MeSH terms or equivalent terminology such as Emtree according to database) and free text words related to the following keywords in English: residents, working hour restrictions, patient safety, resident education, resident well-being, randomized controlled trial, non-randomized controlled trial and pre-post intervention studies. We will search PubMed, MEDLINE, EMBASE, Cochrane Library, Clinicaltrials.gov and Global Index Medicus. There will be no restriction in language or full-text availability.

### Search Strategy

Detailed search strategies developed for each database can be found in the supplementary appendix.

### Study records

#### Data management

The literature search will be run in each database and results will be exported from the database and imported to citation management software (Zotero or similar). These citation files will be organized by folders according to the database of origin and outcome evaluated. After duplicate items are removed, the citation files will then be uploaded to a reference management software such as Rayyan QCRI or similar in order to store articles, facilitate the remote screening process of titles and abstracts among the members of the research team and allow reviewers to mark their decision.

#### Selection process

Two research members will carry out the selection process independently based on the title and abstract of each article to identify pertinent ones to our research purpose. They will screen according to the eligibility criteria outlined above. The reviewer will need to specify the motive to exclude a certain publication. Afterwards, the two independent reviewers will screen the full text of identified articles to confirm the final selection. Any discrepancies on eligibility will be resolved by consulting a third research collaborator. In order to document the article screening process, we will construct a PRISMA flow of information through the different phases of the selection process.

#### Data collection process

Data extraction will be carried out by two independent reviewers by filling out a Google Forms questionnaire that includes data items detailed below (study details, participant characteristics, details of intervention and outcome measures). This information will then be exported to a Microsoft Excel spreadsheet for processing. Any discrepancies in data collection will be resolved with discussion between the two reviewers and, if necessary, consulting with the third author.

## Data items

The extracted data will include:

● Publication information: First author, corresponding author, article title, country, publication year, language
● Population: Sample size (number of participants in each group/arm), inclusion and exclusion criteria, randomization process (if applicable), medical specialty, average age, year of training/rank (intern resident, senior resident)
● Interventions: type of intervention (duty hour restrictions or night float), shift length (number of consecutive duty hours), duration of intervention
● Outcome measures: specify duration of follow-up after intervention and moment at which outcomes were measured
  ○ Primary <u>Patient safety parameters</u>
    ■ 30-day mortality
    ■ 7-day readmission
    ■ 30-day readmission rate
    ■ Adverse events
    ■ Medical errors: if applicable further classified into:
      ● Serious medical error
      ● Near miss
      ● Error with no potential for harm
    ■ In-hospital complications
    ■ Length of stay
    ■ Patient safety indicators
  ○ Secondary <u>Resident Education</u>
    ■ Resident’s examinations scores:
      ● Performance on oral and written American Boards examinations
      ● Performance on in-training examinations
    ■ Resident’s self-reported satisfaction with the educational experience:
      ● Rounds/teaching conference attendance
      ● Independent reading/ self-directed learning
      ● Miss OR/leave during operation
      ● Resident autonomy in medical decision making
      ● Continuity of care
      ● Bedside teaching
      ● Teaching satisfaction
      <u>Resident Well-Being</u>
    ■ Resident’s self-reported levels of rest, professional satisfaction, overall well-being, personal safety, fatigue and work/life balance, which include:
      ● Professional satisfaction
        ○ Job
        ○ Working hours/schedule
        ○ Hands-off
      ● Personal safety
        ○ Percutaneous/Attentional injuries
        ○ sleepiness
        ○ burnout
        ○ number of vehicle crashes
        ○ number of near vehicle crashes
      ● Work/life balance:
        ○ Time for family/friends
        ○ Time for hobbies
        ○ Vacation
        ○ General Health
        ○ Bodily pain
        ○ Mental Health

As previously mentioned, in the case of studies that evaluate working-hour restrictions in healthcare professionals other than medical residents they will only be included if the results are divided and distinguished by occupation. Only results for our relevant population will be reported. If data from a certain trial for a given outcome measure was reported more than once, we will only select the one with the most detailed data.

### Outcomes and prioritization

The belief that extended working hours are detrimental to patient safety is what historically led to resident work hour reforms, but this is currently a controversial topic, which is why patient safety will be set as our primary outcome. However, other variables must be considered when shaping policies regarding medical training such as residents’ sense of well-being and educational achievements, which is why these will be explored as secondary outcomes.

For the primary outcome of interest, we will identify and list all known measurable outcomes including hospital indicators, instruments or scales. If the study reports a type of measurement for our given outcomes not previously identified, it will be included if aligned with our research purposes. For the secondary outcomes of interest, we will apply scales or instruments if available. If not, we will include and document the self-reported subjective responses of participants.

### Risk of bias in individual studies

We will employ the Cochrane risk-of-bias tool for randomized controlled trials (RoB 2) to assess the risk of bias in these types of studies. RoB 2 focuses on several aspects of trial design, conduct and reporting, with questions for each section. The answers to these questions will help identify any potential risk of bias from each domain via an algorithm. After assessing each category, studies will be rated as “high risk”, “low risk”, or “some concerns” (21). For non-randomized studies that include an intervention, the research team will use Cochrane risk of bias in ron-randomized studies of interventions (ROBINS-I). ROBINS-I also contains a series of questions that will assess any arising risk of bias and further classified studies into the following categories: “low risk”, “moderate risk”, “serious risk”, “critical risk” (22). If there is insufficient detailed information to be assessed using RoB 2 or ROBINS-I, the research team will attempt to contact original study investigators to clarify the specific issues. However, if this is not possible, the study will be classified as unclear and will be excluded from the study. Classification of the studies will be conducted independently by two review authors based on the criteria for judging the risk of bias. Disagreements will be resolved by discussing them with a third author for arbitration. Investigators will create a graphic representation of potential bias using Review Manager 5.1 software (RevMan 5.1). Each item will have its own risk of bias assessment independently, and we will avoid using an overall score.

### Strategy for data synthesis

For primary outcomes, we will test for heterogeneity of the included studies by employing the I^2^ statistical test; if considerable heterogeneity is detected (I^2^ ≥ 75%), only qualitative synthesis will be performed. On the contrary, if studies are homogeneous, they will be included in meta-analyses using RevMan 5.1. This will follow a random effects analysis model and the summary outcome measures reported will vary according to the type of data reported. For continuous data, we will calculate the mean difference or standardized mean difference (if the studies employ measurement scales that are not directly comparable to one another). For dichotomous data, the risk ratio (RR) will be calculated. We will then produce a Forest Plot to visualize the results of the meta-analysis. To check for bias, a Funnel plot will be generated and an Egger test will be used. If applicable, the same procedure will be followed for secondary outcomes. Additionally, a qualitative narrative synthesis will be employed to summarize the key findings, population, and methodology of studies using text and tables for both primary and secondary outcomes.

### Quality of evidence

If applicable, the quality of evidence for the primary and secondary outcomes will be assessed through the Grading of Recommendations Assessment, Development and Evaluation (GRADE) system, which rating domains consist of the risk of bias, precision, consistency, indirectness, and publication bias. After applying the GRADE, the quality of evidence will be rated as high, moderate, low, or very low.

## ETHICS AND DISSEMINATION

This systematic review does not require approval by the ethics committee or informed consent. We will request an exoneration from the University’s ethics committee as we will be collecting and synthesizing only published data. Results will be published.

## TIMELINE

| ACTIVITIES | May | Jun | Jul | Aug | Sep | Oct |
| --- | --- | --- | --- | --- | --- | --- |
| Protocol development | X | X |  |  |  |  |
| IRB submission |  |  | X |  |  |  |
| IRB approval |  |  |  | X |  |  |
| Launch database search |  |  |  | X |  |  |
| Abstract & titles screening |  |  |  | X |  |  |
| Full-text screening |  |  |  |  | X |  |
| Development of data extraction template |  |  |  |  | X |  |
| Run pilot of data collection |  |  |  |  | X |  |
| Risk of bias assessment |  |  |  |  | X |  |
| Data entry |  |  |  |  | X |  |
| Data analysis |  |  |  |  | X |  |
| Review results |  |  |  |  | X |  |
| Manuscript drafting |  |  |  |  |  | X |
| Finalize manuscript |  |  |  |  |  | X |

## BUDGET

| ROLE / ITEM | RATE | QUANTITY | METRIC | COST |
| --- | --- | --- | --- | --- |
| **Cochrane Library** | **0** | 1 | subscription | \$0 |
| **Embase** | **0** | 1 | subscription | \$0 |
| **Zotero** | **0** | 1 | subscription | \$0 |
| **Rayyan** | **0** | 1 | subscription | \$0 |
| **Zoom** | **0** | 1 | subscription | \$0 |
| **Office 365** | **\$99** | 1 | subscription | \$99 |
| **Review Manager 5.1** | **0** | 1 | subscription | \$0 |
| **TOTAL** | | | | **\$99** |

## Data Availability

None. Data is not available in other databases.

## REFERENCES

1. Philibert I, Friedmann P, Williams WT. New requirements for resident duty hours. Journal of the American Medical Association. 2002;288(9):1112–4.

2. Nasca TJ, Day SH, Amis ES. The New Recommendations on Duty Hours from the ACGME Task Force. New England Journal of Medicine. 2010;363(2):e3.

3. Summary of Changes to ACGME Common Program Requirements Section VI [Internet]. [cited 2021 Jun 21]. Available from: https://www.acgme.org/What-We-Do/Accreditation/Common-Program-Requirements/Summary-of-Proposed-Changesto-ACGME-Common-Program-Requirements-Section-VI/

4. Ulmer C, Wolman DM, Johns MME. Resident Duty Hours: Enhancing Sleep, Supervision, and Safety Committee on Optimizing Graduate Medical Trainee (Resident) Hours and Work Schedule to Improve Patient Safety, National Research Council [Internet]. 2009 [cited 2021 Jun 21]. Available from: http://www.nap.edu/catalog.php?record_id=12508

5. Lockley SW, Cronin JW, Evans EE, Cade BE, Lee CJ, Landrigan CP, et al. Effect of Reducing Interns’ Weekly Work Hours on Sleep and Attentional Failures. New England Journal of Medicine [Internet]. 2004 Oct 28 [cited 2021 Jun 21];351(18):1829–37. Available from: www.nejm.org

6. Landrigan CP, Rothschild JM, Cronin JW, Kaushal R, Burdick E, Katz JT, et al. Effect of Reducing Interns’ Work Hours on Serious Medical Errors in Intensive Care Units. New England Journal of Medicine [Internet]. 2004 Oct 28 [cited 2021 Jun 21];351(18):1838–48. Available from: www.nejm.org

7. Vidyarthi AR, Arora V, Schnipper JL, Wall SD, Wachter RM. Managing discontinuity in academic medical centers: strategies for a safe and effective resident sign-out. Journal of hospital medicine (Online) [Internet]. 2006 [cited 2021 Jun 21];1(4):257–66. Available from: https://pubmed.ncbi.nlm.nih.gov/17219508/

8. Moura FS, Moura M, Anderson M, de Novais P. Physicians’ working time restriction and its impact on patient safety: an integrative review A restrição da jornada de trabalho do médico e seu impacto na segurança do paciente: uma revisão integrativa. Rev Bras Med Trab. 2018;16(4):482–92.

9. Bolster L, Rourke L. The Effect of Restricting Residents’ Duty Hours on Patient Safety, Resident Well-Being, and Resident Education: An Updated Systematic Review. [cited 2021 Jun 21]; Available from: http://dx.doi.org/10.4300/JGME-D-1400612.1

10. Bilimoria KY, Chung JW, Hedges L v., Dahlke AR, Love R, Cohen ME, et al. National Cluster-Randomized Trial of Duty-Hour Flexibility in Surgical Training. New England Journal of Medicine [Internet]. 2016 Feb 25 [cited 2021 Jun 21];374(8):713–27. Available from: https://www.nejm.org/doi/full/10.1056/NEJMoa1515724

11. Silber JH, Bellini LM, Shea JA, Desai S v., Dinges DF, Basner M, et al. Patient Safety Outcomes under Flexible and Standard Resident Duty-Hour Rules. New England Journal of Medicine [Internet]. 2019 Mar 7 [cited 2021 Jun 21];380(10):905–14. Available from: https://www.nejm.org/doi/full/10.1056/nejmoa1810642

12. Landrigan CP, Rahman SA, Sullivan JP, Vittinghoff E, Barger LK, Sanderson AL, et al. Effect on Patient Safety of a Resident Physician Schedule without 24-Hour Shifts. New England Journal of Medicine [Internet]. 2020 Jun 25 [cited 2021 Jun 21];382(26):2514–23. Available from: https://www.nejm.org/doi/full/10.1056/NEJMoa1900669

13. Mariños A, Otero M, Málaga G, Tomateo J. Coexistencia de síndrome de Burnout y síntomas depresivos en médicos residentes: Estudio descriptivo transversal en un hospital nacional de Lima. Revista Medica Herediana [Internet]. 2011 [cited 2021 Jun 21];22(4):159–60. Available from: http://www.scielo.org.pe/scielo.php?script=sci_arttext&pid=S1018130X2011000400003&lng=es&nrm=iso&tlng=es

14. Inga-Berrospi F, Carlos I, Toro-Huamanchumo J, Lizbeth II, Sanchez A, Torres-Vigo V, et al. Characteristics of the medical residence of teaching hospitals in Lima, Peru. Educación Médica Superior [Internet]. 2016 Jun [cited 2021 Jun 21];30(2). Available from: http://scielo.sld.cuhttp://scielo.sld.cu

15. Mini E, Medina J, Peralta V, Rojas L, Butron J, Gutiérrez EL. Programa de Residentado Médico: Percepciones de los Médicos Residentes en hospitales de Lima y Callao. Revista Peruana de Medicina Experimental y Salud Publica [Internet]. 2002 [cited 2021 Jun 21];32(2):303–10. Available from: http://www.scielo.org.pe/scielo.php?script=sci_arttext&pid=S172646342015000200015&lng=es&nrm=iso&tlng=es

16. Moher D, Shamseer L, Clarke M, Ghersi D, Liberatî A, Petticrew M, et al. Preferred reporting items for systematic review and meta-analysis protocols (PRISMA-P) 2015 statement. Systematic Reviews [Internet]. 2015 [cited 2021 Jun 21]; Available from: http://www.crd.york.ac.uk/prospero

17. Page MJ, Mckenzie JE, Bossuyt PM, Boutron I, Hoffmann TC, Mulrow CD, et al. The PRISMA 2020 statement: an updated guideline for reporting systematic reviews. [cited 2021 Jun 21]; Available from: http://dx.doi.org/10.1136/bmj.n71

18. Thomas EJ, Studdert DM, Burstin HR, Orav EJ, Zeena T, Williams EJ, et al. Incidence and types of adverse events and negligent care in Utah and Colorado. Medical Care [Internet]. 2000 [cited 2021 Jun 21];38(3):261–71. Available from: https://pubmed.ncbi.nlm.nih.gov/10718351/

19. Tamuz M, Thomas EJ. Defining and classifying medical error: lessons for patient safety reporting systems. Suite [Internet]. 463. Available from: www.qshc.com

20. AHRQ Quality Indicators™ Patient Safety Indicators [Internet]. 2015 [cited 2021 Jun 14]. Available from: www.qualityindicators.ahrq.gov

21. Eldridge S, Campbell MK, Campbell MJ, Drahota AK, Giraudeau B, Reeves BC, et al. Revised Cochrane risk of bias tool for randomized trials (RoB 2) Additional considerations for cluster-randomized trials (RoB 2 CRT) Cluster-randomized trials in the context of the Risk of Bias tool Bias arising from the randomization process. RiskofBiasInfo [Internet]. 2020;(November). Available from: https://www.riskofbias.info/welcome/rob-2-0-tool/current-version-of-rob-2

22. Hinneburg I. ROBINS-I: A tool for assessing risk of bias in non-randomised studies of interventions. Medizinische Monatsschrift fur Pharmazeuten. 2017;40(4):175–7.

