## Appendix 1 for "The Effect of Residents’ Working-Hour Restrictions on Patient Safety, Resident Well-Being, and Resident Education: Protocol for a Systematic Review and Meta-Analysis"

**SUPPLEMENTARY APPENDIX**

**Appendix 1. Preliminary search strategies**

A. **Pubmed**

**Descriptor 1: Resident physician**

MeSH terms

“Internship and Residency”[MeSH Terms] OR “Education, Medical, Graduate”[MeSH Terms]

Free terms
“Resident”[tw] OR “junior doctor”[tw] OR “junior physician”[tw] OR “intern”[tw] OR “senior resident”[tw] OR “resident doctor”[tw] OR “resident physician”[tw] OR “medical resident”[tw] OR “surgery resident”[tw] OR “surgical resident”[tw]

**Descriptor 2: Working hours**

MeSH terms

“Personnel Staffing and Scheduling"[MeSH Terms] OR “Workload”[MeSH Terms]

Free terms
“Work hour*”[tw] OR “doctor hour*”[tw] OR “physician hour*”[tw] OR “duty hour*”[tw] OR “resident hour*”[tw] OR “shift hour*”[tw] OR “workload”[tw] OR “work schedule”[tw] OR “night float”[tw] OR “working hour*”[tw] OR “on-call hour*”[tw]

**Descriptor 3: Patient safety**

MeSH terms
"Patient Safety"[MeSH Terms] OR "Medical Errors"[MeSH Terms] OR "Accident Prevention"[MeSH Terms] OR "Patient Harm"[MeSH Terms] OR "Diagnostic Errors"[MeSH Terms] OR "Missed Diagnosis"[MeSH Terms] OR "Medication Errors"[MeSH Terms] OR "Inappropriate Prescribing"[MeSH Terms] OR "Near Miss, Healthcare"[MeSH Terms] OR "Malpractice"[MeSH Terms] OR "Physician Impairment"[MeSH Terms] OR “Hospital mortality”[MeSH Terms] OR “Iatrogenic disease”[MeSH Terms] OR “Quality of healthcare”[MeSH Terms] OR “Patient care”[MeSH Terms] OR “Outcome assessment, healthcare”[MeSH Terms] OR “Treatment outcome”[MeSH Terms] OR “Patient handoff”[MeSH Terms]

Free terms
“serious medical error*”[tw] OR “length of stay”[tw] OR “readmission*”[tw] OR “adverse event*”[tw] OR “patient safety indicator*”[tw] OR “postoperative complication*”[tw] OR “In-Hospital Complication*”[tw]

**Descriptor 4: Resident well-being**

MeSH terms

“Work schedule tolerance”[MeSH Terms] OR “Sleep disorders, circadian rhythm”[MeSH Terms] OR “Stress, psychological”[MeSH Terms] OR “Burnout, professional”[MeSH Terms] OR “Self report”[MeSH Terms] OR “Fatigue”[MeSH Terms] OR “Sleep deprivation”[MeSH Terms] OR “Personal satisfaction”[MeSH Terms] OR “Quality of life”[MeSH Terms] OR “Job satisfaction”[MeSH Terms]

Free terms
“Professional satisfaction” OR “Personal safety” OR “General Health” OR “Mental health” OR “Work and life balance” OR “Time for family” OR “Time for friends” OR “Time for hobbies” OR “Vacation”

**Descriptor 5: Resident education**

MeSH terms
“Education, Medical, Graduate”[MeSH Terms] OR “Professional competence”[MeSH Terms] OR “Clinical competence”[MeSH Terms] OR “Educational measurement”[MeSH Terms] OR “Learning”[MeSH Terms] OR “Resident education”[MeSH Terms] OR “Postgraduate education”[MeSH Terms] OR “Surgical training”[MeSH Terms] OR “Educational measurement”[MeSH Terms]

Free terms
“Postgraduate education”[tw] OR “Surgical training”[tw] OR “Training”[tw] or “Staff training”[tw] or “Residency education”[tw] or “Clinical training”[tw] OR “Clinical skill*”[tw] OR “Round*”[tw] OR “Teaching conference*”[tw] OR “Independent reading”[tw] OR “Self-directed learning”[tw] OR “Miss Or”[tw] OR “Surgical hour*”[tw] OR “Resident autonomy”[tw] OR “Medical decision making”[tw] OR “Bedside teaching”[tw] OR “Teaching satisfaction”[tw] OR “Board examinations”[tw] OR “ABSITE”[tw]

**Descriptor 6: Exclude Nurses**

NOT
“Nursing”[MeSH Terms] OR “Nurses”[MeSH Terms] OR “Nursing staff”[MeSH Terms] OR “Nursing care”[MeSH Terms] OR “Nursing stations”[MeSH Terms] OR “Nursing research”[MeSH Terms] OR “Students, Nursing”[MeSH Terms] OR “Societies, Nursing”[MeSH Terms] OR “Nursing staff”[MeSH Terms] OR “Nursing faculty practice”[MeSH Terms] OR “Nursing services”[MeSH Terms] OR “Nursing process”[MeSH Terms] OR “Faculty, Nursing”[MeSH Terms] OR  “Education, Nursing”[MeSH Terms]

**Descriptor 7: Randomized Controlled Trials (RCTs)**

MeSH terms
"Clinical Trial" [Publication Type] OR "Clinical Trials as Topic"[MeSH Terms] OR "Controlled Clinical Trial" [Publication Type] OR "Pragmatic Clinical Trial" [Publication Type] OR "Clinical Trial, Phase IV" [Publication Type] OR "Adaptive Clinical Trial" [Publication Type] OR "Adaptive Clinical Trials as Topic"[MeSH Terms] OR "Pragmatic Clinical Trials as Topic"[MeSH Terms] OR "Randomized Controlled Trial" [Publication Type]

“Randomised controlled stud*”[tw] OR “Randomised controlled trial*”[tw] OR “RCT”[tw] OR “Randomized controlled trial*”[tw] OR “controlled clinical trial*”[tw] OR “control group stud*”[tw] OR “control group trial*”[tw] OR “controlled trial*”[tw] OR “experimental stud*”[tw]

**Descriptor 8: Non-RCT**

“Quasiexperimental stud*”[tw] OR “quasi-experimental stud*”[tw] OR “mixed method stud*”[tw] OR “pre and post intervention*”[tw] OR “pre and post intervention survey*”[tw] OR “pre and post intervention interview*”[tw]

**Final search A (Descriptor 1 + 2 + 3 + 6 + 7)**
Results: 49

(("Internship and Residency"[MeSH Terms] OR "education, medical, graduate"[MeSH Terms] OR "Resident"[Text Word] OR "junior doctor"[Text Word] OR "junior physician"[Text Word] OR "intern"[Text Word] OR "senior resident"[Text Word] OR "resident doctor"[Text Word] OR "resident physician"[Text Word] OR "medical resident"[Text Word] OR "surgery resident"[Text Word] OR "surgical resident"[Text Word]) AND ("Personnel Staffing and Scheduling"[MeSH Terms] OR "Workload"[MeSH Terms] OR "work hour*"[Text Word] OR "doctor hour*"[Text Word] OR "physician hour*"[Text Word] OR "duty hour*"[Text Word] OR "resident hour*"[Text Word] OR "shift hour*"[Text Word] OR "Workload"[Text Word] OR "work schedule"[Text Word] OR "night float"[Text Word] OR "working hour*"[Text Word] OR "on call hour*"[Text Word]) AND ("Patient Safety"[MeSH Terms] OR "Medical Errors"[MeSH Terms] OR "Accident Prevention"[MeSH Terms] OR "Patient Harm"[MeSH Terms] OR "Diagnostic Errors"[MeSH Terms] OR "Missed Diagnosis"[MeSH Terms] OR "Medication Errors"[MeSH Terms] OR "Inappropriate Prescribing"[MeSH Terms] OR "near miss, healthcare"[MeSH Terms] OR "Malpractice"[MeSH Terms] OR "Physician Impairment"[MeSH Terms] OR "Hospital mortality"[MeSH Terms] OR "Iatrogenic disease"[MeSH Terms] OR "Patient care"[MeSH Terms] OR "Treatment outcome"[MeSH Terms] OR "Patient handoff"[MeSH Terms] OR "serious medical error*"[Text Word] OR "length of stay"[Text Word] OR "readmission*"[Text Word] OR "adverse event*"[Text Word] OR "patient safety indicator*"[Text Word] OR "postoperative complication*"[Text Word] OR "in hospital complication*"[Text Word]) AND ((("Clinical Trial"[Publication Type] OR "Clinical Trials as Topic"[MeSH Terms] OR "Controlled Clinical Trial"[Publication Type] OR "Pragmatic Clinical Trial"[Publication Type] OR "clinical trial, phase iv"[Publication Type] OR "Adaptive Clinical Trial"[Publication Type] OR "Adaptive Clinical Trials as Topic"[MeSH Terms] OR "Pragmatic Clinical Trials as Topic"[MeSH Terms] OR "Randomized Controlled Trial"[Publication Type]) AND "randomised controlled stud*"[Text Word]) OR "randomised controlled trial*"[Text Word] OR "RCT"[Text Word] OR "randomized controlled trial*"[Text Word] OR "controlled clinical trial*"[Text Word] OR "control group stud*"[Text Word] OR "control group trial*"[Text Word] OR "controlled trial*"[Text Word] OR "experimental stud*"[Text Word])) NOT ("Nursing"[MeSH Terms] OR "Nurses"[MeSH Terms] OR "Nursing staff"[MeSH Terms] OR "Nursing care"[MeSH Terms] OR "Nursing stations"[MeSH Terms] OR "Nursing research"[MeSH Terms] OR "students, nursing"[MeSH Terms] OR "societies, nursing"[MeSH Terms] OR "Nursing staff"[MeSH Terms] OR "Nursing faculty practice"[MeSH Terms] OR "Nursing services"[MeSH Terms] OR "Nursing process"[MeSH Terms] OR "faculty, nursing"[MeSH Terms] OR "education, nursing"[MeSH Terms])

**Final search B (Descriptor 1 + 2 + 4 + 6 + 7 + 8)**

Results: 60
(("Internship and Residency"[MeSH Terms] OR "education, medical, graduate"[MeSH Terms] OR "Resident"[Text Word] OR "junior doctor"[Text Word] OR "junior physician"[Text Word] OR "intern"[Text Word] OR "senior resident"[Text Word] OR "resident doctor"[Text Word] OR "resident physician"[Text Word] OR "medical resident"[Text Word] OR "surgery resident"[Text Word] OR "surgical resident"[Text Word]) AND ("Personnel Staffing and Scheduling"[MeSH Terms] OR "Workload"[MeSH Terms] OR "work hour*"[Text Word] OR "doctor hour*"[Text Word] OR "physician hour*"[Text Word] OR "duty hour*"[Text Word] OR "resident hour*"[Text Word] OR "shift hour*"[Text Word] OR "Workload"[Text Word] OR "work schedule"[Text Word] OR "night float"[Text Word] OR "working hour*"[Text Word] OR "on call hour*"[Text Word]) AND ("Work schedule tolerance"[MeSH Terms] OR "sleep disorders, circadian rhythm"[MeSH Terms] OR "stress, psychological"[MeSH Terms] OR "burnout, professional"[MeSH Terms] OR "Self report"[MeSH Terms] OR "Fatigue"[MeSH Terms] OR "Sleep deprivation"[MeSH Terms] OR "Personal satisfaction"[MeSH Terms] OR "Quality of life"[MeSH Terms] OR "Job satisfaction"[MeSH Terms] OR "Professional satisfaction"[All Fields] OR "Personal safety"[All Fields] OR "General Health"[All Fields] OR "Mental health"[All Fields] OR "Work and life balance"[All Fields] OR (("time"[MeSH Terms] OR "time"[All Fields]) AND ("familialities"[All Fields] OR "familiality"[All Fields] OR "familially"[All Fields] OR "familials"[All Fields] OR "familie"[All Fields] OR "family"[MeSH Terms] OR "family"[All Fields] OR "familial"[All Fields] OR "families"[All Fields] OR "family s"[All Fields] OR "familys"[All Fields])) OR (("time"[MeSH Terms] OR "time"[All Fields]) AND ("friend s"[All Fields] OR "friending"[All Fields] OR "friends"[MeSH Terms] OR "friends"[All Fields] OR "friend"[All Fields])) OR (("time"[MeSH Terms] OR "time"[All Fields]) AND ("hobbies"[MeSH Terms] OR "hobbies"[All Fields] OR "hobby"[All Fields])) OR "Vacation"[All Fields]) AND ("Clinical Trial"[Publication Type] OR "Clinical Trials as Topic"[MeSH Terms] OR "Controlled Clinical Trial"[Publication Type] OR "Pragmatic Clinical Trial"[Publication Type] OR "clinical trial, phase iv"[Publication Type] OR "Adaptive Clinical Trial"[Publication Type] OR "Adaptive Clinical Trials as Topic"[MeSH Terms] OR "Pragmatic Clinical Trials as Topic"[MeSH Terms] OR "Randomized Controlled Trial"[Publication Type] OR "randomised controlled stud*"[Text Word] OR "randomised controlled trial*"[Text Word] OR "RCT"[Text Word] OR "randomized controlled trial*"[Text Word] OR "controlled clinical trial*"[Text Word] OR "control group stud*"[Text Word] OR "control group trial*"[Text Word] OR "controlled trial*"[Text Word] OR "experimental stud*"[Text Word] OR "quasiexperimental stud*"[Text Word] OR "quasi experimental stud*"[Text Word] OR "mixed method stud*"[Text Word] OR "pre and post intervention*"[Text Word] OR "pre and post intervention survey*"[Text Word])) NOT ("Nursing"[MeSH Terms] OR "Nurses"[MeSH Terms] OR "Nursing staff"[MeSH Terms] OR "Nursing care"[MeSH Terms] OR "Nursing stations"[MeSH Terms] OR "Nursing research"[MeSH Terms] OR "students, nursing"[MeSH Terms] OR "societies, nursing"[MeSH Terms] OR "Nursing staff"[MeSH Terms] OR "Nursing faculty practice"[MeSH Terms] OR "Nursing services"[MeSH Terms] OR "Nursing process"[MeSH Terms] OR "faculty, nursing"[MeSH Terms] OR "education, nursing"[MeSH Terms])

**Final search C (Descriptor 1 + 2 + 5 + 6 + 7 + 8**)
Results: 116
((("Internship and Residency"[MeSH Terms] OR "education, medical, graduate"[MeSH Terms] OR "Resident"[Text Word] OR "junior doctor"[Text Word] OR "junior physician"[Text Word] OR "intern"[Text Word] OR "senior resident"[Text Word] OR "resident doctor"[Text Word] OR "resident physician"[Text Word] OR "medical

resident"[Text Word] OR "surgery resident"[Text Word] OR "surgical resident"[Text Word]) AND ("Personnel Staffing and Scheduling"[MeSH Terms] OR "Workload"[MeSH Terms] OR "work hour*"[Text Word] OR "doctor hour*"[Text Word] OR "physician hour*"[Text Word] OR "duty hour*"[Text Word] OR "resident hour*"[Text Word] OR "shift hour*"[Text Word] OR "Workload"[Text Word] OR "work schedule"[Text Word] OR "night float"[Text Word] OR "working hour*"[Text Word] OR "on call hour*"[Text Word]) AND ("Clinical Trial"[Publication Type] OR "Clinical Trials as Topic"[MeSH Terms] OR "Controlled Clinical Trial"[Publication Type] OR "Pragmatic Clinical Trial"[Publication Type] OR "clinical trial, phase iv"[Publication Type] OR "Adaptive Clinical Trial"[Publication Type] OR "Adaptive Clinical Trials as Topic"[MeSH Terms] OR "Pragmatic Clinical Trials as Topic"[MeSH Terms] OR "Randomized Controlled Trial"[Publication Type] OR "randomised controlled stud*"[Text Word] OR "randomised controlled trial*"[Text Word] OR "RCT"[Text Word] OR "randomized controlled trial*"[Text Word] OR "controlled clinical trial*"[Text Word] OR "control group stud*"[Text Word] OR "control group trial*"[Text Word] OR "controlled trial*"[Text Word] OR "experimental stud*"[Text Word] OR "quasiexperimental stud*"[Text Word] OR "quasi experimental stud*"[Text Word] OR "mixed method stud*"[Text Word] OR "pre and post intervention*"[Text Word] OR "pre and post intervention survey*"[Text Word])) NOT ("Nursing"[MeSH Terms] OR "Nurses"[MeSH Terms] OR "Nursing staff"[MeSH Terms] OR "Nursing care"[MeSH Terms] OR "Nursing stations"[MeSH Terms] OR "Nursing research"[MeSH Terms] OR "students, nursing"[MeSH Terms] OR "societies, nursing"[MeSH Terms] OR "Nursing staff"[MeSH Terms] OR "Nursing faculty practice"[MeSH Terms] OR "Nursing services"[MeSH Terms] OR "Nursing process"[MeSH Terms] OR "faculty, nursing"[MeSH Terms] OR "education, nursing"[MeSH Terms])) AND ("education, medical, graduate"[MeSH Terms] OR "Professional competence"[MeSH Terms] OR "Clinical competence"[MeSH Terms] OR "Educational measurement"[MeSH Terms] OR "Learning"[MeSH Terms] OR "Educational measurement"[MeSH Terms] OR "Postgraduate education"[Text Word] OR "Surgical training"[Text Word] OR "Training"[Text Word] OR "Staff training"[Text Word] OR "Residency education"[Text Word] OR "Clinical training"[Text Word] OR "clinical skill*"[Text Word] OR "round*"[Text Word] OR "teaching conference*"[Text Word] OR "Independent reading"[Text Word] OR "Self-directed learning"[Text Word] OR "surgical hour*"[Text Word] OR "Resident autonomy"[Text Word] OR "Medical decision making"[Text Word] OR "Bedside teaching"[Text Word] OR "Teaching satisfaction"[Text Word] OR "Board examinations"[Text Word] OR "ABSITE"[Text Word])

**Global Index Medicus**

**Descriptor 1: Resident physician**

MeSH terms

mh:(Internship and Residency) OR mh:(Education, Medical, Graduate)

Free terms
tw:(resident*) OR tw:("junior doctor*") OR tw:("junior physician*") OR tw:(intern*) OR tw:("senior resident*") OR tw:("resident doctor*") OR tw:("resident physician*") OR tw:("medical resident*") OR tw:("surgery resident*") OR tw:("surgical resident*") OR tw:("surgical trainee*")

**Descriptor 2: Working hours**

MeSH terms

mh:(Personnel Staffing and Scheduling) OR mh:(Workload)

Free terms
tw:(“Work hour*”) OR tw:(“doctor hour*”) OR tw:(“physician hour*”) OR tw:(“duty hour”) OR tw:(“resident hour*”) OR tw:(“shift hour*”) OR tw:(workload) OR tw:(“work schedule”) OR tw:(“night float”) OR tw:(working hour*) OR tw:(“on-call hour*”) OR tw:(working time restriction) OR tw:(flexible schedul*)

**Descriptor 3: Exclude Nurses**
NOT
mh:(Nursing) OR mh:(Nurses) OR mh:(Nursing staff) OR mh:(Nursing care) OR mh:(Nursing stations) OR mh:(Nursing research) OR mh:(Students, Nursing) OR mh:(Societies, Nursing) OR mh:(Nursing staff) OR mh:(Nursing faculty practice) OR mh:(Nursing services) OR mh:(Nursing process) OR mh:(Faculty, Nursing) OR mh:(Education, Nursing) OR tw:(nursing) OR tw:(nursing staff) OR tw:(nurs*)

**Final search** (Descriptor 1 + Descriptor 2 + Descriptor 3)
Results: 553

tw:((tw:(mh:(internship AND residency) OR mh:(education, medical, graduate) OR tw:(resident*) OR tw:(“junior doctor*”) OR tw:(“junior physician*”) OR tw:(intern*) OR tw:(“senior resident*”) OR tw:(“resident doctor*”) OR tw:(“resident physician*”) OR tw:(“medical resident*”) OR tw:(“surgery resident*”) OR tw:(“surgical resident*”) OR tw:(“surgical trainee*”))) AND (tw:(mh:(personnel staffing AND scheduling) OR mh:(workload) OR tw:(“work hour*”) OR tw:(“doctor hour*”) OR tw:(“physician hour*”) OR tw:(“duty hour”) OR tw:(“resident hour*”) OR tw:(“shift hour*”) OR tw:(workload) OR tw:(“work schedule”) OR tw:(“night float”) OR tw:(working hour*) OR tw:(“on-call hour*”) OR tw:(working time restriction) OR tw:(flexible schedul*))) AND NOT (tw:(mh:(nursing) OR mh:(nurses) OR mh:(nursing staff) OR mh:(nursing care) OR mh:(nursing stations) OR mh:(nursing research) OR mh:(students, nursing) OR mh:(societies, nursing) OR mh:(nursing staff) OR mh:(nursing faculty practice) OR mh:(nursing services) OR mh:(nursing process) OR mh:(faculty, nursing) OR mh:(education, nursing) OR tw:(nursing) OR tw:(nursing staff) OR tw:(nurs*))))

### <u>Cochrane</u>

**Descriptor 1: Resident physician**
#1 MeSH descriptor: [Internship and Residency] explode all trees 1284
#2 MeSH descriptor: [Physicians] explode all trees 2117

**Descriptor 2: Working hours**
#3 MeSH descriptor: [Workload] explode all trees 400
#4 MeSH descriptor: [Work Schedule Tolerance] explode all trees 155
#5 MeSH descriptor: [Personnel Staffing and Scheduling] explode all trees 624

**Final search (Descriptor 1 + Descriptor 2)**
Results: 76
#6 (#1 OR #2) AND (#3 OR #4 OR #5) 76

**Embase**

**Descriptor 1: Resident physician**

Emtree Term
'internship'/exp OR 'resident'/exp

Embase field code
‘Resident’:ti,ab,kw,de OR ‘junior doctor’:ti,ab,kw,de OR ‘junior physician’:ti,ab,kw,de OR ‘intern’:ti,ab,kw,de OR ‘senior resident’:ti,ab,kw,de OR ‘resident doctor’:ti,ab,kw,de OR ‘resident physician’:ti,ab,kw,de OR ‘medical resident’:ti,ab,kw,de OR ‘surgery resident’:ti,ab,kw,de OR ‘surgical resident’:ti,ab,kw,de

**Descriptor 2: Working hours**

Emtree Term
'health care personnel management'/exp OR 'workload'/exp

Embase field code
‘Work hour*’:ti,ab,kw,de OR ‘doctor hour*’:ti,ab,kw,de OR ‘physician hour*’:ti,ab,kw,de OR ‘duty hour*’:ti,ab,kw,de OR ‘resident hour*’:ti,ab,kw,de OR ‘shift hour*’:ti,ab,kw,de OR ‘workload’:ti,ab,kw,de OR ‘work schedule’:ti,ab,kw,de OR ‘night float’:ti,ab,kw,de OR ‘working hour*’:ti,ab,kw,de OR ‘on-call hour*’:ti,ab,kw,de

**Descriptor 3: Patient safety**

Emtree terms
'patient safety'/exp OR 'patient safety indicator'/exp OR 'medical error'/exp OR 'accident prevention'/exp OR 'patient harm'/exp OR 'diagnostic error'/exp OR 'medication error'/exp OR 'inappropriate prescribing'/exp OR 'near miss (health care)'/exp OR 'malpractice'/exp OR 'hospital mortality'/exp OR 'iatrogenic disease'/exp OR 'health care quality'/exp OR 'patient care'/exp OR 'outcome assessment'/exp OR 'treatment outcome'/exp OR 'clinical handover'/exp

Embase field code
‘serious medical error*’:ti,ab,kw,de OR ‘length of stay’:ti,ab,kw,de OR ‘readmission*’:ti,ab,kw,de OR ‘adverse event*’:ti,ab,kw,de OR ‘patient safety indicator*’:ti,ab,kw,de OR ‘postoperative complication*’:ti,ab,kw,de OR ‘In-Hospital Complication*’:ti,ab,kw,de

**Descriptor 4: Resident well-being**

Emtree terms
'work schedule'/exp OR 'sleep disorder'/exp OR 'circadian rhythm sleep disorder'/exp OR 'physiological stress'/exp OR 'professional burnout'/exp OR 'self report'/exp OR 'fatigue'/exp OR 'sleep deprivation'/exp OR 'satisfaction'/exp OR 'quality of life'/exp OR 'job satisfaction'/exp

Embase field code

‘Professional satisfaction’:ti,ab,kw,de OR ‘Personal safety’:ti,ab,kw,de OR ‘General Health’:ti,ab,kw,de OR ‘Mental health’:ti,ab,kw,de OR ‘Work and life balance’:ti,ab,kw,de OR ‘Time for family’:ti,ab,kw,de OR ‘Time for friends’:ti,ab,kw,de OR ‘Time for hobbies’:ti,ab,kw,de OR ‘Vacation’:ti,ab,kw,de

**Descriptor 5: Resident education**

Emtree terms
'medical education'/exp OR 'medical education'/exp OR 'clinical competence'/exp OR 'education'/exp OR 'learning'/exp OR 'postgraduate education'/exp OR 'surgical training'/exp

Embase field code
‘Postgraduate education’:ti,ab,kw,de OR ‘Surgical training’:ti,ab,kw,de OR ‘Training’:ti,ab,kw,de or ‘Staff training’:ti,ab,kw,de or ‘Residency education’:ti,ab,kw,de or ‘Clinical training’:ti,ab,kw,de OR ‘Clinical skill*’:ti,ab,kw,de OR ‘Round*’:ti,ab,kw,de OR ‘Teaching conference*’:ti,ab,kw,de OR ‘Independent reading’:ti,ab,kw,de OR ‘Self-directed learning’:ti,ab,kw,de OR ‘Miss Or’:ti,ab,kw,de OR ‘Surgical hour*’:ti,ab,kw,de OR ‘Resident autonomy’:ti,ab,kw,de OR ‘Medical decision making’:ti,ab,kw,de OR ‘Bedside teaching’:ti,ab,kw,de OR ‘Teaching satisfaction’:ti,ab,kw,de OR ‘Board examinations’:ti,ab,kw,de OR ‘ABSITE’:ti,ab,kw,de

**Descriptor 6: Exclude Nurses**

NOT “
'nurse'/exp OR 'nursing'/exp OR 'nursing staff'/exp OR 'nursing care'/exp OR 'nursing station'/exp OR 'nursing research'/exp OR 'nursing students'/exp OR 'nursing organization'/exp OR 'nursing education'/exp OR 'nursing education'/exp

**Descriptor 7: Randomized Controlled Trials (RCTs) and non-RCT**

Emtree terms
'clinical trial'/exp OR 'controlled clinical trial'/exp OR 'randomized controlled trial'/exp OR 'pragmatic trial'/exp OR 'phase 4 clinical trial'/exp OR 'adaptive clinical trial'/exp

Embase field code
‘Randomised controlled stud*’:ti,ab,kw,de OR ‘Randomised controlled trial*’:ti,ab,kw,de OR ‘RCT’:ti,ab,kw,de OR ‘Randomized controlled trial*’:ti,ab,kw,de OR ‘controlled clinical trial*’:ti,ab,kw,de OR ‘control group stud*’:ti,ab,kw,de OR ‘control group trial*’:ti,ab,kw,de OR ‘controlled trial*’:ti,ab,kw,de OR ‘experimental stud*’:ti,ab,kw,de

**Descriptor 8: non-RCT**
Embase field code
‘quasiexperimental stud*’:ti,ab,kw,de OR ‘quasi-experimental stud*’:ti,ab,kw,de OR ‘mixed method stud*’:ti,ab,kw,de OR ‘pre and post intervention*’:ti,ab,kw,de OR ‘pre and post intervention survey*’:ti,ab,kw,de OR ‘pre and post intervention interview*’:ti,ab,kw,de

**Final search A**
Descriptor 1 + descriptor 2 + descriptor 3 + descriptor 7 - descriptor 6

Results: 116

('internship'/exp OR 'internship' OR 'resident'/exp OR 'resident' OR 'resident':ti,ab,kw,de OR 'junior doctor':ti,ab,kw,de OR 'junior physician':ti,ab,kw,de OR 'intern':ti,ab,kw,de OR 'senior resident':ti,ab,kw,de OR 'resident doctor':ti,ab,kw,de OR 'resident physician':ti,ab,kw,de OR 'medical resident':ti,ab,kw,de OR 'surgery resident':ti,ab,kw,de OR 'surgical resident':ti,ab,kw,de) AND ('health care personnel management'/exp OR 'health care personnel management' OR 'workload'/exp OR 'workload' OR 'work hour*':ti,ab,kw,de OR 'doctor hour*':ti,ab,kw,de OR 'physician hour*':ti,ab,kw,de OR 'duty hour*':ti,ab,kw,de OR 'resident hour*':ti,ab,kw,de OR 'shift hour*':ti,ab,kw,de OR 'workload':ti,ab,kw,de OR 'work schedule':ti,ab,kw,de OR 'night float':ti,ab,kw,de OR 'working hour*':ti,ab,kw,de OR 'on-call hour*':ti,ab,kw,de) AND ('patient safety'/exp OR 'patient safety' OR 'patient safety indicator'/exp OR 'patient safety indicator' OR 'medical error'/exp OR 'medical error' OR 'accident prevention'/exp OR 'accident prevention' OR 'patient harm'/exp OR 'patient harm' OR 'diagnostic error'/exp OR 'diagnostic error' OR 'medication error'/exp OR 'medication error' OR 'inappropriate prescribing'/exp OR 'inappropriate prescribing' OR 'near miss (health care)'/exp OR 'near miss (health care)' OR 'malpractice'/exp OR 'malpractice' OR 'hospital mortality'/exp OR 'hospital mortality' OR 'iatrogenic disease'/exp OR 'iatrogenic disease' OR 'health care quality'/exp OR 'health care quality' OR 'patient care'/exp OR 'patient care' OR 'outcome assessment'/exp OR 'outcome assessment' OR 'treatment outcome'/exp OR 'treatment outcome' OR 'clinical handover'/exp OR 'clinical handover' OR 'serious medical error*':ti,ab,kw,de OR 'length of stay':ti,ab,kw,de OR 'readmission*':ti,ab,kw,de OR 'adverse event*':ti,ab,kw,de OR 'patient safety indicator*':ti,ab,kw,de OR 'postoperative complication*':ti,ab,kw,de OR 'in-hospital complication*':ti,ab,kw,de) AND ('clinical trial'/exp OR 'clinical trial' OR 'controlled clinical trial'/exp OR 'controlled clinical trial' OR 'randomized controlled trial'/exp OR 'randomized controlled trial' OR 'pragmatic trial'/exp OR 'pragmatic trial' OR 'phase 4 clinical trial'/exp OR 'phase 4 clinical trial' OR 'adaptive clinical trial'/exp OR 'adaptive clinical trial' OR 'randomised controlled stud*':ti,ab,kw,de OR 'randomised controlled trial*':ti,ab,kw,de OR 'rct':ti,ab,kw,de OR 'randomized controlled trial*':ti,ab,kw,de OR 'controlled clinical trial*':ti,ab,kw,de OR 'control group stud*':ti,ab,kw,de OR 'control group trial*':ti,ab,kw,de OR 'controlled trial*':ti,ab,kw,de OR 'experimental stud*':ti,ab,kw,de) NOT ('nurse'/exp OR 'nurse' OR 'nursing'/exp OR 'nursing' OR 'nursing staff'/exp OR 'nursing staff' OR 'nursing care'/exp OR 'nursing care' OR 'nursing station'/exp OR 'nursing station' OR 'nursing research'/exp OR 'nursing research' OR 'nursing students'/exp OR 'nursing students' OR 'nursing organization'/exp OR 'nursing organization' OR 'nursing education'/exp OR 'nursing education')

**Final search B**

Descriptor 1 + descriptor 2 + descriptor 4 + descriptor 7 + descriptor 8 - descriptor 6

Results: 144

('internship'/exp OR 'resident'/exp OR 'resident':ti,ab,kw,de OR 'junior doctor':ti,ab,kw,de OR 'junior physician':ti,ab,kw,de OR 'intern':ti,ab,kw,de OR 'senior resident':ti,ab,kw,de OR 'resident doctor':ti,ab,kw,de OR 'resident physician':ti,ab,kw,de OR 'medical resident':ti,ab,kw,de OR 'surgery resident':ti,ab,kw,de OR 'surgical resident':ti,ab,kw,de) AND ('health care personnel management'/exp OR 'workload'/exp OR 'work hour*':ti,ab,kw,de OR 'doctor hour*':ti,ab,kw,de OR 'physician hour*':ti,ab,kw,de OR 'duty hour*':ti,ab,kw,de OR 'resident hour*':ti,ab,kw,de OR 'shift hour*':ti,ab,kw,de OR 'workload':ti,ab,kw,de OR 'work schedule':ti,ab,kw,de OR 'night float':ti,ab,kw,de OR 'working hour*':ti,ab,kw,de OR 'on-call hour*':ti,ab,kw,de) AND ('medical education'/exp OR 'clinical competence'/exp OR 'education'/exp OR 'learning'/exp OR 'postgraduate education'/exp OR 'surgical training'/exp

OR 'postgraduate education':ti,ab,kw,de OR 'surgical training':ti,ab,kw,de OR 'training':ti,ab,kw,de OR 'staff training':ti,ab,kw,de OR 'residency education':ti,ab,kw,de OR 'clinical training':ti,ab,kw,de OR 'clinical skill*':ti,ab,kw,de OR 'round*':ti,ab,kw,de OR 'teaching conference*':ti,ab,kw,de OR 'independent reading':ti,ab,kw,de OR 'self-directed learning':ti,ab,kw,de OR 'miss or':ti,ab,kw,de OR 'surgical hour*':ti,ab,kw,de OR 'resident autonomy':ti,ab,kw,de OR 'medical decision making':ti,ab,kw,de OR 'bedside teaching':ti,ab,kw,de OR 'teaching satisfaction':ti,ab,kw,de OR 'board examinations':ti,ab,kw,de OR 'absite':ti,ab,kw,de) AND ('clinical trial'/exp OR 'controlled clinical trial'/exp OR 'randomized controlled trial'/exp OR 'pragmatic trial'/exp OR 'phase 4 clinical trial'/exp OR 'adaptive clinical trial'/exp OR 'randomised controlled stud*':ti,ab,kw,de OR 'randomised controlled trial*':ti,ab,kw,de OR 'rct':ti,ab,kw,de OR 'randomized controlled trial*':ti,ab,kw,de OR 'controlled clinical trial*':ti,ab,kw,de OR 'control group stud*':ti,ab,kw,de OR 'control group trial*':ti,ab,kw,de OR 'controlled trial*':ti,ab,kw,de OR 'experimental stud*':ti,ab,kw,de OR 'quasiexperimental stud*':ti,ab,kw,de OR 'quasi-experimental stud*':ti,ab,kw,de OR 'mixed method stud*':ti,ab,kw,de OR 'pre and post intervention*':ti,ab,kw,de OR 'pre and post intervention survey*':ti,ab,kw,de OR 'pre and post intervention interview*':ti,ab,kw,de) NOT ('nurse'/exp OR 'nursing'/exp OR 'nursing staff'/exp OR 'nursing care'/exp OR 'nursing station'/exp OR 'nursing research'/exp OR 'nursing students'/exp OR 'nursing organization'/exp OR 'nursing education'/exp)

**Final search C**

<u>Descriptor 1 + descriptor 2 + descriptor 5 + descriptor 7 + descriptor 8 - descriptor 6</u>

Results: 86

('internship'/exp OR 'resident'/exp OR 'resident':ti,ab,kw,de OR 'junior doctor':ti,ab,kw,de OR 'junior physician':ti,ab,kw,de OR 'intern':ti,ab,kw,de OR 'senior resident':ti,ab,kw,de OR 'resident doctor':ti,ab,kw,de OR 'resident physician':ti,ab,kw,de OR 'medical resident':ti,ab,kw,de OR 'surgery resident':ti,ab,kw,de OR 'surgical resident':ti,ab,kw,de) AND ('health care personnel management'/exp OR 'workload'/exp OR 'work hour*':ti,ab,kw,de OR 'doctor hour*':ti,ab,kw,de OR 'physician hour*':ti,ab,kw,de OR 'duty hour*':ti,ab,kw,de OR 'resident hour*':ti,ab,kw,de OR 'shift hour*':ti,ab,kw,de OR 'workload':ti,ab,kw,de OR 'work schedule':ti,ab,kw,de OR 'night float':ti,ab,kw,de OR 'working hour*':ti,ab,kw,de OR 'on-call hour*':ti,ab,kw,de) AND ('work schedule'/exp OR 'sleep disorder'/exp OR 'circadian rhythm sleep disorder'/exp OR 'physiological stress'/exp OR 'professional burnout'/exp OR 'self report'/exp OR 'fatigue'/exp OR 'sleep deprivation'/exp OR 'satisfaction'/exp OR 'quality of life'/exp OR 'job satisfaction'/exp OR 'professional satisfaction':ti,ab,kw,de OR 'personal safety':ti,ab,kw,de OR 'general health':ti,ab,kw,de OR 'mental health':ti,ab,kw,de OR 'work and life balance':ti,ab,kw,de OR 'time for family':ti,ab,kw,de OR 'time for friends':ti,ab,kw,de OR 'time for hobbies':ti,ab,kw,de OR 'vacation':ti,ab,kw,de) AND ('clinical trial'/exp OR 'controlled clinical trial'/exp OR 'randomized controlled trial'/exp OR 'pragmatic trial'/exp OR 'phase 4 clinical trial'/exp OR 'adaptive clinical trial'/exp OR 'randomised controlled stud*':ti,ab,kw,de OR 'randomised controlled trial*':ti,ab,kw,de OR 'rct':ti,ab,kw,de OR 'randomized controlled trial*':ti,ab,kw,de OR 'controlled clinical trial*':ti,ab,kw,de OR 'control group stud*':ti,ab,kw,de OR 'control group trial*':ti,ab,kw,de OR 'controlled trial*':ti,ab,kw,de OR 'experimental stud*':ti,ab,kw,de OR 'quasiexperimental stud*':ti,ab,kw,de OR 'quasi-experimental stud*':ti,ab,kw,de OR 'mixed method stud*':ti,ab,kw,de OR 'pre and post intervention*':ti,ab,kw,de OR 'pre and post intervention survey*':ti,ab,kw,de OR 'pre and post intervention interview*':ti,ab,kw,de) NOT ('nurse'/exp OR 'nursing'/exp OR 'nursing staff'/exp OR 'nursing care'/exp OR 'nursing station'/exp OR 'nursing research'/exp OR 'nursing students'/exp OR 'nursing organization'/exp OR 'nursing education'/exp)

**ClinicalTrials.gov**

Keywords

- Resident work hours
- Duty hours
- Work hour reduction
- Resident scheduling
- Night float
- Residents working
- Resident wellness and burnout
- Patient safety
- Resident education

**Final search**

Results: 12

AREA[ConditionSearch] ("resident work hours" OR "duty hours" OR "work hour reduction" OR "resident scheduling" OR "night float" OR "residents working" OR "resident wellness and burnout" OR "patient safety and resident education")
